# Evaluating the Harmonization of Native Digital and Digitized ECGs for ECG-AI Research

**DOI:** 10.64898/2026.09.21.26363586

**Authors:** Elsayed Z. Soliman, Hassaan A. Bukhari, Luke Patterson, Semseddin Moldibi, Ibrahim Karabayir, Oguz Akbilgic

**Author notes:** **<u>Corresponding Author:</u>** Elsayed Z. Soliman MD, MSc, MS, Epidemiological Cardiology Research Center (EPICARE), Wake Forest School of Medicine, Medical Center Blvd, Winston-Salem, NC 27157.

## Abstract

Large epidemiologic studies have historical electrocardiogram (ECG) collections limited to paper tracings or scanned images rather than native digital files, limiting ECG-AI applications. We compared 15 ECGs available as native digital XML files and as PDF tracings digitized using ECGScan software. Both versions were processed using an ensemble of five convolutional neural networks predicting 10-year heart failure risk. Predictions were strongly correlated (Pearson *r*=0.804; Spearman ρ=0.893). Predicted probabilities were (mean ± SD) 0.195 ± 0.048 for native digital ECGs and 0.171 ± 0.042 for digitized ECGs. These preliminary findings support further evaluation of digitized ECGs for ECG-AI applications.

## Introduction

Large epidemiologic studies frequently include ECGs acquired across different eras and platforms. Earlier examinations may be preserved only as paper tracings or scanned images, whereas later visits are often stored as native digital signals. As previously described by Waits and Soliman, converting paper ECGs into analyzable digital waveforms could preserve valuable historical data and enable automated measurements, standardized classification, and contemporary computational analyses, although signal recovery and validation remain important challenges [1].

Recent digitization methods have demonstrated high correlation between reconstructed and native ECG waveforms, including ECGs subjected to printing and scanning [2,3]. However, waveform similarity does not necessarily establish that native digital and digitized ECGs will generate comparable outputs when analyzed by an established electrocardiographic artificial intelligence (ECG-AI) model. This question is particularly relevant when combining studies with different ECG-storage formats or evaluating longitudinal cohorts in which earlier visits used paper ECGs and later visits used native digital acquisition.

We therefore compared ECG-AI outputs from native digital ECGs with those from digitized versions of the same recordings. A previously developed and validated heart-failure risk prediction model [4] was used as an analytic tool to assess downstream harmonization rather than to evaluate heart-failure prediction itself.

## Methods

Fifteen deidentified resting 12-lead ECGs were obtained from the EPICARE database at Wake Forest School of Medicine, Winston-Salem, North Carolina. Each ECG had originally been acquired digitally and was evaluated in two formats: (1) the native digital export in GE MUSE XML format, sampled at 500 Hz for 10 seconds; and (2) a digitized waveform derived from a PDF rendering of the same ECG, used to simulate the digitization of a paper tracing. The PDF was rasterized at 600 dots per inch and digitized at standard calibration settings of 25 mm/s and 10 mm/mV using ECGScan software (AMPS LLC, New York, NY, USA). The digitized pixel traces were converted to microvolts using the recorded calibration and resampled to 500 Hz.

Both ECG versions were processed identically using the previously developed deep residual convolutional neural network (CNN) architecture for prediction of 10-year heart failure risk [4]. The first second of each recording was excluded to allow signal stabilization, and the remaining nine seconds (4500 samples across 12 leads) were submitted to each of five independently trained models (0–4). Their predicted probabilities were averaged to generate one ensemble output for each ECG version. The ensemble approach improves generalization and reduces prediction variance compared with predictions from a single model. The heart-failure model was used as an analytic probe to determine whether digitization preserved information relevant to a downstream ECG-AI application.

Predictions from the native digital and digitized ECGs were compared using Pearson and Spearman correlation coefficients. Paired differences were calculated as digitized minus native digital output. The mean paired difference, mean absolute paired difference, and range of paired differences were calculated to characterize the magnitude and direction of disagreement between formats. Agreement was further examined using Bland–Altman analysis, with 95% limits of agreement calculated as the mean paired difference ±1.96 standard deviations of the paired differences. All statistical tests were two-sided, and all analyses were performed in Python using SciPy.

## Results

Fifteen deidentified 12-lead ECGs were available in both native digital XML and PDF-derived digitized XML formats. Representative paired tracings appeared similar on visual inspection (Figure 1A). The mean ensemble ECG-AI output was 0.195 for native digital ECGs and 0.171 for digitized ECGs. Outputs were strongly correlated between formats (Pearson r = 0.804; Spearman ρ = 0.893; both P <0.001; Figure 1B). Digitized ECGs yielded lower outputs on average, with a mean paired difference of −0.024 and 95% limits of agreement of −0.080 to 0.032 (Table 1; Figure 1C). The mean absolute paired difference was 0.031, and individual paired differences ranged from −0.063 to 0.035.

**Figure 1.**
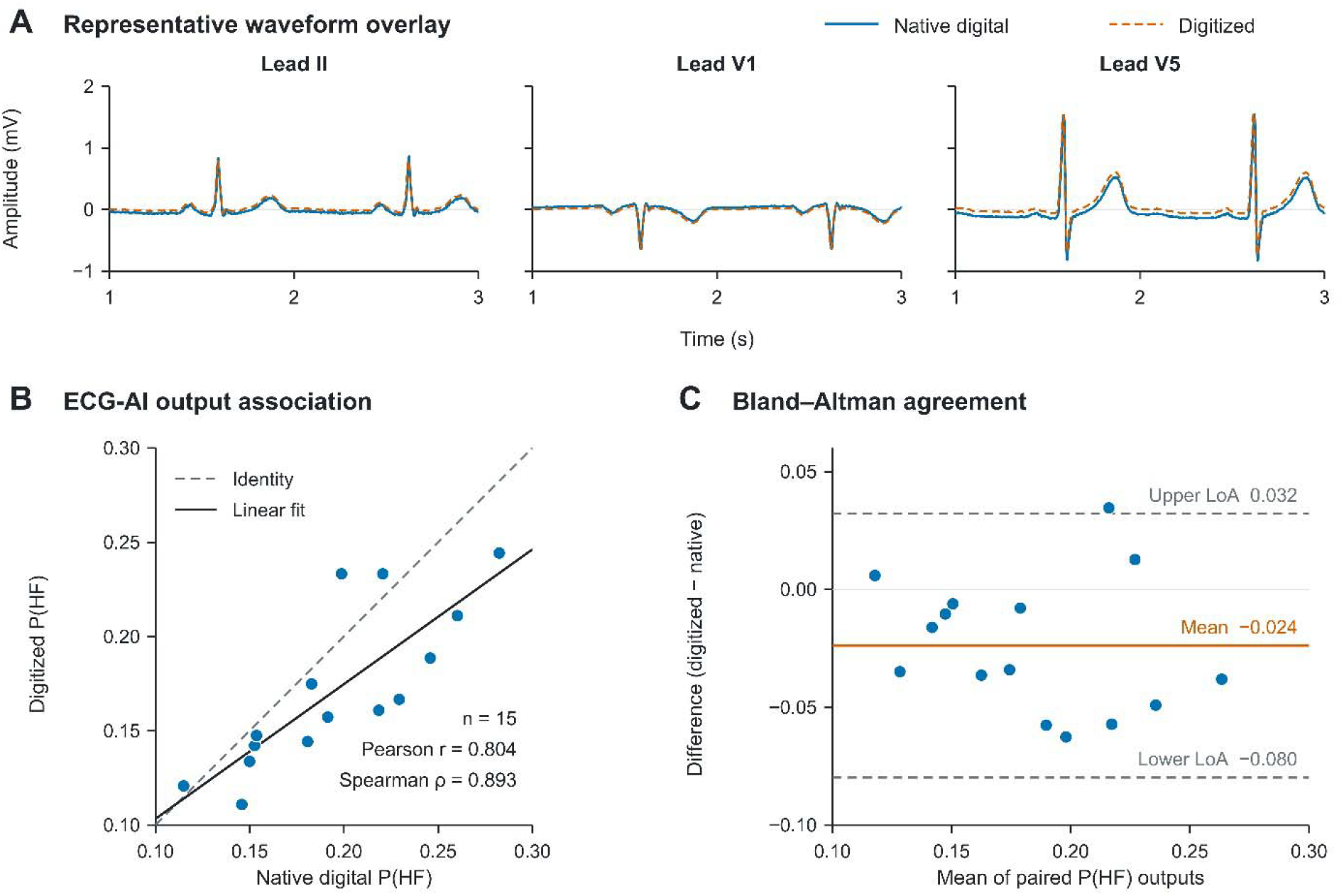
Comparison of native digital and digitized ECGs. (A) Overlaid native digital and PDF-derived digitized waveforms from leads II, V1, and V5, showing a representative two-second segment (1–3 seconds). (B) Relationship between ensemble ECG-AI outputs from the two formats. The dashed diagonal represents the line of identity, and the solid line represents the fitted linear regression. (C) Bland–Altman plot showing digitized minus native digital outputs against their paired means. The solid horizontal line represents the mean difference, and the dashed lines represent the 95% limits of agreement. P(HF), model-predicted probability of heart failure; LoA, limits of agreement.

**Table 1.** Agreement between native digital and digitized ECG-AI outputs.

| **Measure** | **Result** |
| --- | --- |
| Number of paired ECGs | 15 |
| Native digital output, mean ± SD | 0.195 ± 0.048 |
| Digitized output, mean ± SD | 0.171 ± 0.042 |
| Mean paired difference, digitized minus native digital | -0.024 |
| Mean absolute paired difference | 0.031 |
| Range of paired differences | -0.063 to 0.035 |
| Pearson correlation | 0.804 |
| Spearman correlation | 0.893 |
| Bland–Altman 95% limits of agreement | -0.080 to 0.032 |

## Discussion

In this proof-of-concept study, ECG-AI outputs derived from digitized ECGs were strongly correlated with outputs from the corresponding native digital recordings. The particularly high rank correlation suggests that the relative ordering of individuals by the model was largely preserved after digitization. These findings support the feasibility of harmonizing ECGs stored in different formats for research applications in which earlier examinations are available only as paper tracings or images, whereas later examinations or external validation cohorts contain native digital ECGs.

Prior studies have primarily evaluated digitization by comparing reconstructed waveforms with their native digital signals. Wu et al. reported high waveform correlations after ECGs were printed, scanned, and redigitized, while Demolder et al. demonstrated generally high signal fidelity across ECG images with varying quality and acquisition conditions [2,3]. Our study extends this work by evaluating digitization at the level of a downstream ECG-AI output. This application-level assessment is important because small waveform differences that appear acceptable using conventional signal-similarity measures may still influence a neural network prediction.

Nevertheless, the digitized ECGs yielded slightly lower predicted probabilities on average. Thus, strong correlation should not be interpreted as complete agreement or interchangeability between formats. When native digital and digitized ECGs are combined, particularly across study visits, acquisition format may need to be considered during model calibration, sensitivity analyses, or statistical adjustment. The principal value of harmonization may be the ability to apply a common analytic pipeline across historical and contemporary ECG collections, thereby expanding sample size, extending longitudinal follow-up, and enabling external validation in cohorts for which native digital signals are unavailable.

This study has several limitations. It included only 15 ECGs, represented a relatively narrow range of model predictions, and evaluated a single digitization workflow and one ECG-AI application. Moreover, the PDFs were generated from originally digital ECGs and therefore did not reproduce artifacts encountered with aging, folding, printing, or scanning of actual paper tracings. Clinical discrimination, calibration, and threshold-based classification could not be evaluated. Accordingly, these findings establish preliminary feasibility rather than equivalence. Larger studies using authentic scanned paper ECGs, different layouts and image qualities, and multiple ECG-AI models are needed before native digital and digitized ECGs can be used interchangeably.

## Data Availability

All data produced in the present study are available upon reasonable request to the authors

## Acknowledgment

The authors thank AMPS LLC for providing access to the ECGScan software and technical assistance with the ECG digitization process. AMPS LLC had no role in the study design, statistical analysis, interpretation of the findings, or preparation of the manuscript.

